# Mining medical narratives on geriatric falls to predict post-fall hospitalization via survival model and language models

**DOI:** 10.1101/2025.10.15.25336949

**Authors:** Lisa Y.W. Tang

## Abstract

Timely admission to the emergency department is a crucial determinant of patient outcomes. Conversely, unnecessary hospital admissions can overburden health systems and induce anxiety and stress among patients, their families, and care-givers. This study examines these implications in the geriatric population by investigating the hypothesis that delay time, i.e. the interval between injury and hospital admission, is associated with patient outcomes post admission. As delay times are not typically captured in electronic medical records, we leverage a large database from an open challenge where short narratives describing the patient injuries and treatment were made publicly available. Accordingly, we developed prognostic survival models based on large-language models that predict time to an adverse outcome using features extracted from the textual narratives, as well as additional data provided by the challenge, e.g. data about patients’ baseline characteristics, conditions of their injuries. To this end, we found that models incorporating textual embeddings achieved a dynamic area under the curve (D-AUC) of 0.713-0.715, compared to 0.637–0.649 for models without textual features, when evaluated on an external cohort. This study provides preliminary evidence that the textual data collected at the time of patients triage can be useful for patient prognosis. Future studies will examine how the same data, collected right after fall events could be used to project patient’s recovery progress.

## 1 Introduction

Timely admission to the emergency department (ED) is crucial for determining outcomes in elderly patients. Research has shown that delays in ED admission can lead to poorer care, particularly for patients presenting with severe pain and complex medical needs (Pines & Hollander, 2008).

Many studies have reported associations between waiting times in the ED and patient outcomes in the general population (Blunt et al., 2015). In pediatric settings, Guttmann et al. (2011) reported that longer waiting times in the ED are associated with increased short-term mortality and hospital admissions in children. Among older adults, McCusker et al. (2000) found that delays in initial ED visits and hospital admissions are significant predictors of repeat ED visits. Pines & Hollander (2008); Pines et al. (2013) also reported that longer wait times and delays in ED admission are associated with poorer pain management and overall care in seniors. Similarly, Garcia & Gonzalez (2014) observed that delayed treatment worsened health outcomes in elderly patients.

Conversely, unnecessary hospital admissions can overburden health systems, contributing to the so-called “ED overcrowding crisis”, and leading to inefficiencies and increased healthcare costs (Blunt et al., 2015). These unnecessary admissions can also induce significant anxiety and stress among patients, their families, and caregivers, further adding to the existing caregiver burden (Adelman et al., 2014).

The aforementioned findings collectively underscore the critical importance of timely medical treatment through ED admission and hospitalization, as well as accurate severity assessment at the time of injury. While such assessments can be challenging to perform in community-based or resourcelimited settings, they remain essential for avoiding unnecessary hospital visits and mitigating ED overcrowding, which can delay care for patients in genuine need.

Accordingly, this study investigates whether **narrative text collected during oral interviews with elderly patients at the time of a fall contains predictive information about patient outcomes**. Specifically, we test the hypothesis that these short, unstructured narratives, often recorded by medical abstractors or non-clinical personnel, contain clinically meaningful signals that can inform triage or follow-up decisions.

Several prior studies have applied survival models to time-to-event prediction tasks in healthcare, with promising results. However, few studies have explored the use of narrative injury descriptions to predict long-term outcomes in the geriatric population. Our work addresses this gap by extracting predictive features from injury narratives captured at times of hospital visits, and using survival modeling to evaluate the association between delay times and patient prognosis.

By exploring the predictive value of triage narratives, this work contributes to ongoing efforts at the intersection of natural language processing and emergency medicine. While this study does not compare model outputs directly with clinician judgment, it lays the groundwork for future studies exploring how such models could support clinical decision-making. Furthermore, by focusing on a form of data already captured in routine workflows, our findings point toward potential applications in developing assistive tools for early risk stratification in geriatric care.

## 2 Methods

### 2.1 Data

We employ data from a 2023 community-based challenge hosted by DrivenData and originally provisioned by the National Electronic Injury Surveillance System (NEISS) (Shields et al., 2024):

- `PRIMARY` cohort: 115,000 emergency department (ED) visits due to unintentional falls among older adults (aged 65+) between 2019 and 2022;
- `SUPPLEMENTARY` cohort: 418,000 fall-related ED visits spanning 2013 to 2022.

The narrative information was manually prepared by trained medical abstractors following specific coding instructions, namely:

- Age and sex should be included at the beginning of the narrative, e.g., “50YOF” for a 50-year-old female;
- The sequence of events should be described in detail;
- Correct names and spelling of products should be used;
- Clinical diagnoses should be placed at the end, marked with “Dx”;
- Only the first word of each sentence should be capitalized.

Further details are available on the challenge website.

### 2.2 Pre-Processing of the narratives

Our natural language pre-processing (NLP) pipeline consists of a sequence of rule-based transformations, applied uniformly across all narrative texts. No supervised training data was used to optimize or tune these steps.

First, we applied **variable-driven text pruning** to remove elements already captured by structured variables (e.g., sex, age), as well as any content following markers such as “DX”, which often denote diagnostic or metadata fields. Next, we performed **manual typo correction** for frequent and easily recognizable misspellings (e.g., “nite” was corrected to “night”).

We then applied **symbol normalization**, reducing repeated symbols (e.g., ***, >>>) to a single instance or spelling them out (e.g., & to “and”, @ to “at”). **Medical abbreviations** were expanded into their standard English equivalents to improve lexical consistency.

All narratives were subsequently **lemmatized** using the `spaCy` library, with safeguards to preserve context-specific expressions (e.g., “left foot” was not incorrectly reduced to “leave foot”). Finally, we performed **spell checking and word splitting**: if a word was not recognized by the `PySpellChecker` library, it was split into two substrings, provided the first substring was a valid English word.

### 2.3 Extraction of Word Embeddings (*X*)

After data cleaning, we extracted different sets of word embeddings to encode the processed narratives. These included:

- **All-MPNet-Base-V2 (WE1)**, based on the MPNet architecture, designed to capture contextual relationships between tokens (Ashqar & Mutlu, 2023);
- **All-MiniLM-L6-V2 (WE2)**, a compact and efficient variant of BERT that maintains competitive performance while reducing computational cost (Yin & Zhang, 2024; Galli et al., 2024); and
- **Quantized MiniLM (WE3)**, a compact (“quantized”) version of WE2 accessed through LangChain’s `GPT4AllEmbeddings` wrapper. This version is optimized for lightweight, CPU-based inference and trades off some accuracy for efficiency and portability (Anand et al., 2023).

### 2.4 Extraction of Delay Times from Narratives (*y*)

Delay times (*t*), i.e. the time from fall to ED visit that eventually led to an event outcome were not coded in the datasets. In this work, we define the **outcome as hospital admission that eventually led to medical treatment(s)**.

To extract *t* from the narratives, we implemented a keyword-based search strategy, targeting specific phrases and terms that indicated the time elapsed between the fall incident and hospital visit, e.g. “1DAY AGO”, “YESTERDAY”, “LAST NITE”. Cases with indeterminable delay times were excluded from all subsequent analyses. Additionally, patients with dispositions not matching the predefined outcome definitions were treated as censored; this includes those who left the hospital before being seen.

A crucial pre-processing step is to systematically remove words and phrases that explicitly mention delay times from the narratives. This masking process must be completed before computing word embeddings to prevent leakage of delay time information into our predictive models.

### 2.1 MODEL DEVELOPMENT

Model development was performed using the `PRIMARY` cohort, which we partitioned into training (*n* = 5,064), validation (*n* = 5,064), and test (*n* = 212,356) sets. The `SUPPLEMENTARY` cohort used as an external test set spanned multiple years (2013–2020) and was further divided into early and late subsets to assess temporal generalization. Notably, the size of our evaluation data is comparable to publicly available survival datasets, such as SUPPORT (*n* = 9,105) and FLCHAIN (*n* = 7,894), previously used in benchmark studies of clinical time-to-event modeling (Chapfuwa et al., 2020).

We adopted **eXtreme Gradient Boosting (XGBoost)** (Chen & Guestrin, 2016) as our primary survival model due to its flexibility, performance, and scalability for high-dimensional data. XGBoost offers several advantages in this context: it can be adapted to handle right-censored data; it is nonparametric, requiring no assumptions about the distribution of survival times; and it provides interpretable outputs such as feature importance scores. Moreover, its computational efficiency makes it well suited for large-scale datasets, particularly those derived from unstructured text.

As a statistical baseline, we also implemented the **Cox proportional hazards model (CoxNet)**. For hyperparameter tuning of XGBoost, we employed the `Optuna` framework (Akiba et al., 2019), which uses Bayesian optimization to efficiently explore the hyperparameter space. We defined a search space including learning rate, tree depth, and gamma, and ran 100 optimization trials. At each step, `Optuna` proposed hyperparameter configurations based on prior trial performance, using the validation set for feedback. This process enabled us to identify high-performing model configurations and improve generalization to unseen data.

## 3 Results

To evaluate the impact of different embedding strategies on survival prediction, we systematically compared the performance of multiple word embedding models. We developed a suite of survival models, each incorporating no more than three input components: raw embeddings, their dimensionally reduced variants, and optionally, non-textual covariates (sex, age category, location of fall, etc.). This modular design allowed us to isolate the effect of text-derived features and assess their interaction with structured clinical variables.

In the Cox regression analysis, we excluded the variable indicating fire involvement due to its extreme sparsity (which would otherwise lead to unstable parameter estimates and degenerate model behavior). Similarly, we excluded the diagnosis variable, consistent with our preprocessing of the narrative text where “DX” information was stripped prior to embedding. This decision was motivated by the high collinearity between diagnosis codes and patient disposition outcomes.

Table 1 reports the performance of each configuration using the dynamic area-under-curve (D-AUC) (Chapfuwa et al., 2020). As the table summarizes, the XGBoost-based survival models exhibited strong predictive performance across different input configurations. These results highlight the model’s capacity to capture meaningful signals from both textual and structured data sources.

**Table 1:** The D-AUC was obtained using an XGBoost model optimized with the Optuna package. Non-textual covariates include age category, sex, and location of the fall.

| Model | Inputs | Devel. Val. | Devel. Test | Ext. Period 1 | Period 2 |
| --- | --- | --- | --- | --- | --- |
| CoxNet | Non-textual covariates | 0.575 | 0.551 | 0.572 | 0.600 |
| XGBoost | Non-textual covariates | 0.661 | 0.633 | 0.649 | 0.637 |
| XGBoost | WE1 | 0.698 | 0.685 | 0.696 | 0.716 |
|  | WE1 + Non-textual covariates | 0.674 | 0.629 | 0.614 | 0.677 |
|  | WE2 | 0.685 | 0.678 | 0.685 | 0.707 |
|  | WE2 + Non-textual covariates | **0.738** | **0.721** | **0.713** | **0.715** |
|  | WE3 | 0.683 | 0.686 | 0.687 | 0.696 |
|  | WE3 + Non-textual covariates | 0.680 | 0.686 | 0.687 | 0.696 |

## 4 Discussion & Conclusions

XGBoost, the top-performing model, achieved a D-AUC of 0.721 on the internal test set and 0.713–0.715 on the external validation set (see `SUPPLEMENTARY`). To assess the added value of textual narratives, we trained the same model using only non-textual covariates (i.e. age category, sex, and location of the fall). Optimized in the same manner, this variant yielded a substantially lower D-AUC of 0.633 on the internal test set and 0.637–0.649 on the external validation sets. This performance drop underscores the predictive contribution of narrative-based features in modeling patient outcomes and delay times.

This study has several limitations. First, the retrospective nature of the analysis introduces potential selection and documentation bias, as narrative detail may reflect clinician concern rather than underlying patient risk. Second, although language models enable scalable representation of clinical narratives, their outputs remain indirect proxies of clinical concepts and may encode latent biases present in the source text. Our results should therefore be interpreted as a methodological demonstration rather than a deployment-ready model.

Despite these limitations, our findings demonstrate that injury narratives collected at the time of emergency visits encode information highly predictive of patient outcomes and hospitalization delays. Understanding this relationship can inform improvements in emergency care practices, policy development, and caregiver support, ultimately enhancing outcomes and reducing strain on healthcare systems.

## Data Availability

Public may create a free account and subsequently download data through DrivenData.

https://www.drivendata.org/competitions/217/cdc-fall-narratives/

## Acknowledgment

We gratefully acknowledge UBC Advanced Research Computing and the (Digital Research Alliance of Canada) for providing in-kind computational resources and technical support. We also thank Kim Chuen Tang and Tsui Shan Tong for their unwavering support throughout this research.

